# A nationwide multistate analysis estimating the rates and risks of transferring critically ill COVID-19 patients during the Delta and Omicron waves in Germany

**DOI:** 10.1101/2023.03.31.23287964

**Authors:** Matthäus Lottes, Marlon Grodd, Linus Grabenhenrich, Martin Wolkewitz

**Affiliations:** Robert Koch Institute, Department of Methods Development, Research Infrastructure and Information Technology, Berlin, Germany; Institute of Medical Biometry and Statistics, Faculty of Medicine and Medical Center - University of Freiburg, Freiburg, Germany

**Keywords:** Intensive care unit, SARS-CoV-2 variant of concern, inter-hospital transfer, multistate model, competing risk

## Abstract

**Background:** The spread of several SARS-CoV-2 variants of concern (VOC) led to increasing numbers of patients with coronavirus disease 2019 (COVID-19) in German intensive care units (ICU), resulting in capacity shortages and even transfers of COVID-19 ICU patients between federal states in late 2021. Comprehensive evidence on the impact of predominant VOC, in this case Delta and Omicron, on inter-hospital transfers of COVID-19 ICU patients remains scarce.

**Methods:** A retrospective cohort study was conducted from July 01, 2021 until May 31, 2022 using nationwide reimbursement inpatient count data of COVID-19 ICU patients and weekly sequence data of VOC in Germany. A multivariable Poisson regression analysis was performed to estimate incidence rates and incidence rate ratios (IRR) for competing events of transfer, discharge and death, adjusted for VOC infection, age group and sex. For corresponding risk estimation, a multistate model for the clinical trajectory in ICU was applied.

**Results:** Omicron versus Delta infection yielded estimated adjusted IRR of 1.23 (95% CI, 1.16 – 1.30) for transfer, 2.27 (95% CI, 2.20 – 2.34), for discharge and 0.98 (95% CI, 0.94 – 1.02) for death. For death in ICU, estimated adjusted IRR increased progressively with age up to 4.09 (95% CI, 3.74 – 4.47) for those 90 years and older. COVID-19 ICU patients with Omicron infection were at comparatively higher estimated risk of discharge, whereas the estimated risk of transfer and death were higher for those with Delta infection.

**Conclusions:** Inter-hospital transfers and discharges occurred more frequently in COVID-19 ICU patients with Omicron infection than in those with Delta infection, who in turn had a higher estimated risk of death. Age emerges as a relevant determinant for fatal clinical trajectories in COVID-19 ICU patients and imposes close therapeutic care.

## Introduction

Since early 2020, the coronavirus disease 2019 (COVID-19) pandemic has posed major challenges to the German health care system both on structural and individual level [1-5]. In particular, intensive care units (ICU) have been severely impacted during periods of increased COVID-19 caseloads with a total daily occupancy of more than 5,000 adult COVID-19 patients distributed across approximately 1,300 adult ICU nationwide [6]. Among these, the proportion of patients who required invasive ventilation or extracorporeal membrane oxygenation exceeded 60% on several occasions. In terms of mortality, the maximum number of deaths of COVID-19 patients in ICU surpassed 200 per day, cumulating to a total number of over 50,000 deaths of COVID-19 patients in German ICU by the end of May 2022 [7]. Given all these immense challenges for critical care in Germany, efficient resource allocation, including transfers of COVID-19 ICU patients between federal states, has become crucial to maintain health care delivery and avoid system collapse [8-12].

One of the main factors leading to periods of critical capacity shortages in ICU is the spread of several SARS-CoV-2 variants of concern (VOC) in the German population [13, 14]. By the end of 2020, the Delta VOC increased the risk of hospitalization and fatal outcome compared with the previously dominant Alpha VOC (15-19). ICU occupancy and respiratory support subsequently increased [15-19]. ICU occupancy and respiratory support subsequently increased [7, 20]. In contrast, the Omicron VOC, first reported in South Africa and rapidly circulated in Germany in late 2021, led to reduced disease severity, but its sub-lineages demonstrated high transmissibility and immune escape [21-27]. As a result, the number of new infections in the German population has risen sharply, and with it the number of COVID-19 ICU patients, which has even necessitated their nationwide allocation [28, 29]. So far, there is limited comprehensive information on the impact of predominant VOC on critical care and especially on inter-hospital transfers of COVID-19 ICU patients in Germany. Because similar situations might occur in the resource-constraint ICU setting, further evidence is important, in particular to promote efficient patient allocation and to reduce ICU mortality.

Therefore, the objective of our study was to determine the impact of Delta and Omicron VOC on critical care in Germany with a focus on transfers of COVID-19 ICU patients between hospitals. To do so, we aimed to estimate and compare transfer, discharge and death rates in ICU associated with Delta and Omicron infection. We furthermore attempted to estimate the associated risk of transfer, discharge and death by using the aforementioned rates to model the clinical trajectory of COVID-19 ICU patients [30, 31].

## Methods

### Data sources and structure

The data for this retrospective cohort study were provided by the Institute for the Hospital Remuneration System GmbH (InEK) and include the reimbursement data of all hospitals in Germany that are subject to the data transmission obligation [32]. Data consists of nationwide total daily counts of prevalent, transferred, discharged and death COVID-19 cases admitted to a German ICU by age group (0-9, 10-19, …, 90+) and sex (female, male) from July 1, 2021 until May 31, 2022. Since the InEK only provides data on patients who have completed clinical treatment, underreporting of daily counts is to be expected at the end of this observation period, which was therefore limited to April 30, 2022. Based on weekly sequence data published by the Robert Koch Institute (RKI) on the number and proportions of VOC in Germany, the entire analysis was further restricted to calendar periods for Delta VOC from September 13, 2021 (begin of calendar week 37) until November 21, 2021 (end of calendar week 46) and for Omicron VOC from February 14, 2022 (begin of calendar week 7) until April 30, 2022 [13]. Within these calendar periods almost exclusively Delta and Omicron VOC were sequenced, including their respective sub-lineages, and we therefore used them as surrogate measures of infection impact in our analysis. Thus, the sequenced VOC were not considered individually but grouped into Delta and Omicron infection categories, hereafter referred to as VOC infection.

### Basic multistate model

Based on InEK data, a multistate approach (Fig. 1) was used to model the clinical trajectory of COVID-19 patients in ICU and considering the competing risk of transfer, discharge and death [33, 34]. Accordingly, patients begin with ICU admission (state 0). They may remain in this state until they are either transferred to another hospital as part of clinical care (state 1), discharged from ICU (state 2) or die in ICU (state 3). Consequently, COVID-19 patients may also be discharged or die in ICU after an inter-hospital transfer (from state 1 to state 2 or state 3). The multistate model depends on the hazard rates (*α*_01_, *α*_02_, *α*_03_, *α*_12_, *α*_13_) between the states admission, transfer, discharge and death. Given the InEK data structure, these hazard rates depend on VOC infection, age group and sex. However, within each of these categories, the hazard rates were assumed to remain constant over time, discharge and death rates to be independent of inter-hospital transfers (*α*_02_ = *α*_12_, *α*_03_ = *α*_13_) and inter-hospital transfers to occur only once during the clinical treatment of a COVID-19 ICU patient. As just mentioned, we also assumed that discharges (state 2) and deaths (state 3) always occurred in ICU, although this was not unambiguous from the data provided and both events may have also occurred outside the ICU during inpatient follow-up.

**Fig. 1.**
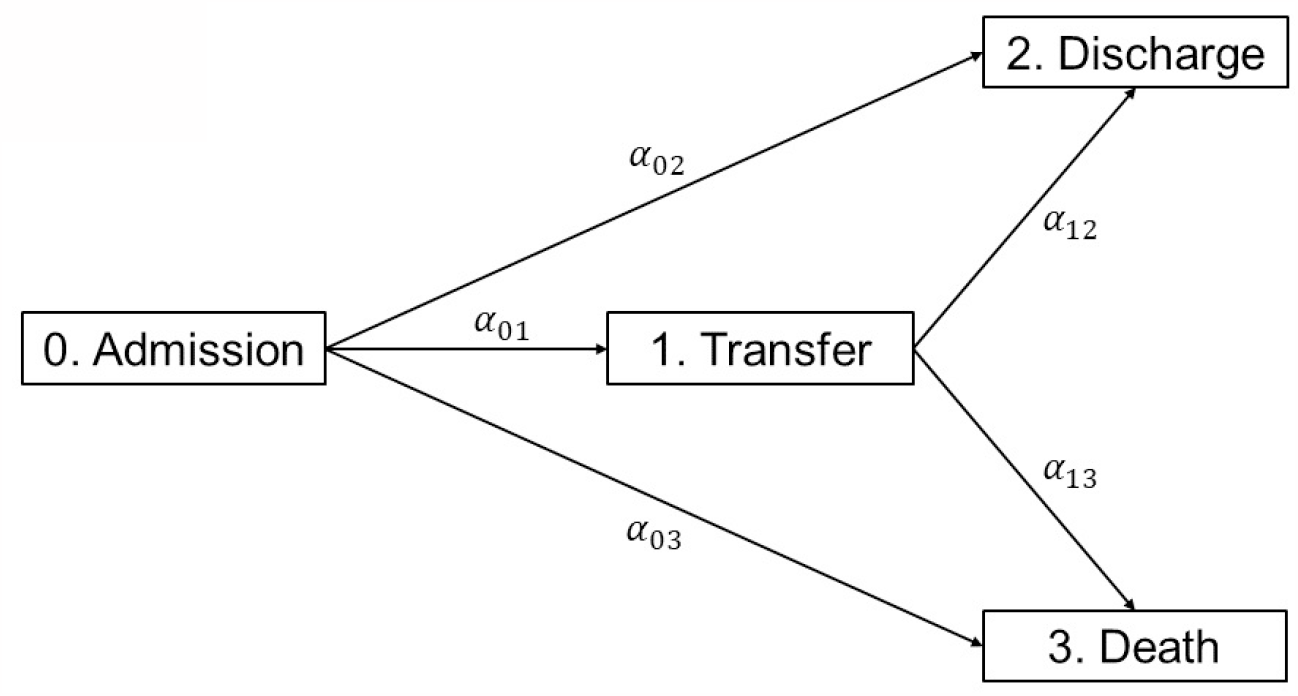
Schematic representation of a multistate model for the clinical trajectory of COVID-19 patients in German intensive care units based on data from the Institute for the Hospital Remuneration System GmbH (InEK)

### Statistical analysis

Based on our assumption of constant hazard rates, a multivariable Poisson regression analysis was performed to estimate the incidence rates and incidence rate ratios (IRR) of transfer, discharge, and death of COVID-19 ICU patients associated with VOC infection. Hence, a separate Poisson regression model was fitted for these outcomes, adjusted for categories of VOC infection, age group and sex of the InEK data (Table 1). Delta was the VOC infection reference due to the timing. The 60-69 age group was chosen as the age reference since it has the highest count of prevalent COVID-19 cases. For sex, female COIVD-19 ICU patients were the reference but without specific reason. For interpretation, an estimated IRR above 1 (below 1) indicates a higher (lower) event rate of transfer, discharge and death in the categories VOC infection, age group and sex compared with their reference.

**Table 1.** Estimated incidence rate ratios with 95% confidence intervals and p-values of transfer, discharge and death of COVID-19 patients in German intensive care units, adjusted for SARS-CoV-2 variant of concern infection, age group and sex

| Predictors | Transfer |  |  | Discharge |  |  | Death |  |  |
| --- | --- | --- | --- | --- | --- | --- | --- | --- | --- |
|  | Incidence Rate Ratio | 95% CI<sup>a</sup> | p<sup>b</sup> | Incidence Rate Ratio | 95% CI<sup>a</sup> | p<sup>b</sup> | Incidence Rate Ratio | 95% CI<sup>a</sup> | p<sup>b</sup> |
| Delta | 1.00 | *Reference* |  | 1.00 | *Reference* |  | 1.00 | *Reference* |  |
| Omicron | 1.23 | 1.16 – 1.30 | <0.001 | 2.27 | 2.20 – 2.34 | <0.001 | 0.98 | 0.94 – 1.02 | 0.298 |
| Age 0-9 | 0.57 | 0.41 – 0.76 | <0.001 | 1.88 | 1.72 – 2.05 | <0.001 | 0.14 | 0.08 – 0.22 | <0.001 |
| Age 10-19 | 1.54 | 1.17 – 1.99 | 0.001 | 2.40 | 2.15 – 2.67 | <0.001 | 0.34 | 0.21 – 0.51 | <0.001 |
| Age 20-29 | 1.54 | 1.27 – 1.84 | <0.001 | 2.33 | 2.15 – 2.53 | <0.001 | 0.30 | 0.22 – 0.41 | <0.001 |
| Age 30-39 | 1.52 | 1.33 – 1.72 | <0.001 | 1.76 | 1.65 – 1.88 | <0.001 | 0.41 | 0.34 – 0.49 | <0.001 |
| Age 40-49 | 1.35 | 1.21 – 1.50 | <0.001 | 1.40 | 1.32 – 1.48 | <0.001 | 0.52 | 0.46 – 0.58 | <0.001 |
| Age 50-59 | 1.19 | 1.09 – 1.30 | <0.001 | 1.17 | 1.12 – 1.23 | <0.001 | 0.70 | 0.65 – 0.76 | <0.001 |
| Age 60-69 | 1.00 | *Reference* |  | 1.00 | *Reference* |  | 1.00 | *Reference* |  |
| Age 70-79 | 0.90 | 0.83 – 0.98 | 0.015 | 1.02 | 0.98 – 1.06 | 0.319 | 1.50 | 1.42 – 1.59 | <0.001 |
| Age 80-89 | 0.98 | 0.90 – 1.07 | 0.640 | 1.31 | 1.26 – 1.37 | <0.001 | 2.58 | 2.44 – 2.72 | <0.001 |
| Age 90+ | 1.18 | 0.99 – 1.41 | 0.065 | 2.01 | 1.87 – 2.16 | <0.001 | 4.09 | 3.74 – 4.47 | <0.001 |
| Female | 1.00 | *Reference* |  | 1.00 | *Reference* |  | 1.00 | *Reference* |  |
| Male | 0.90 | 0.85 – 0.95 | <0.001 | 0.83 | 0.81 – 0.85 | <0.001 | 1.04 | 1.00 – 1.08 | 0.075 |

<sup>a</sup> Confidence interval
<sup>b</sup> P-value

To estimate the risk of transfer, discharge and death of COVID-19 ICU patients associated with VOC infection, we adapted the approach of von Cube et al. [35] and converted the constant hazard rates between the states (Fig. 1) into estimated cumulative probabilities, i.e., risks adjusted for categories of VOC infection, age group and sex dependent on time (day) since ICU admission, noted as *t*:

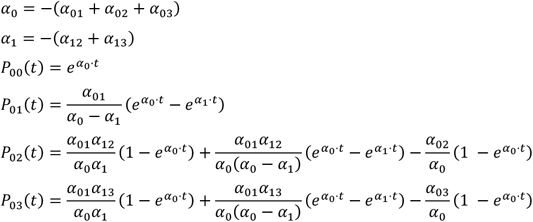

The cumulative probability of being transferred to another hospital was estimated by:

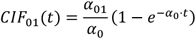

The cumulative probability of being discharged from ICU was estimated by *CIF*_02_(*t*), equal to *P*_02_(*t*), and the cumulative probability to die in ICU was estimated by *CIF*_03_ (*t*), equal to *P*_03_(*t*). In the following, we primarily considered *CIF*_01_(*t*), *CIF*_02_(*t*), *CIF*_03_ (*t*) associated with VOC infection up to day 15 since ICU admission (Fig. 2 – 4), although longer durations of treatment were of course possible, and by age group, as age had already been shown to be a major determinant for COVID-19 related in-hospital mortality [36-38]. All analyses were performed using R version 4.2.2.

**Fig. 2.**
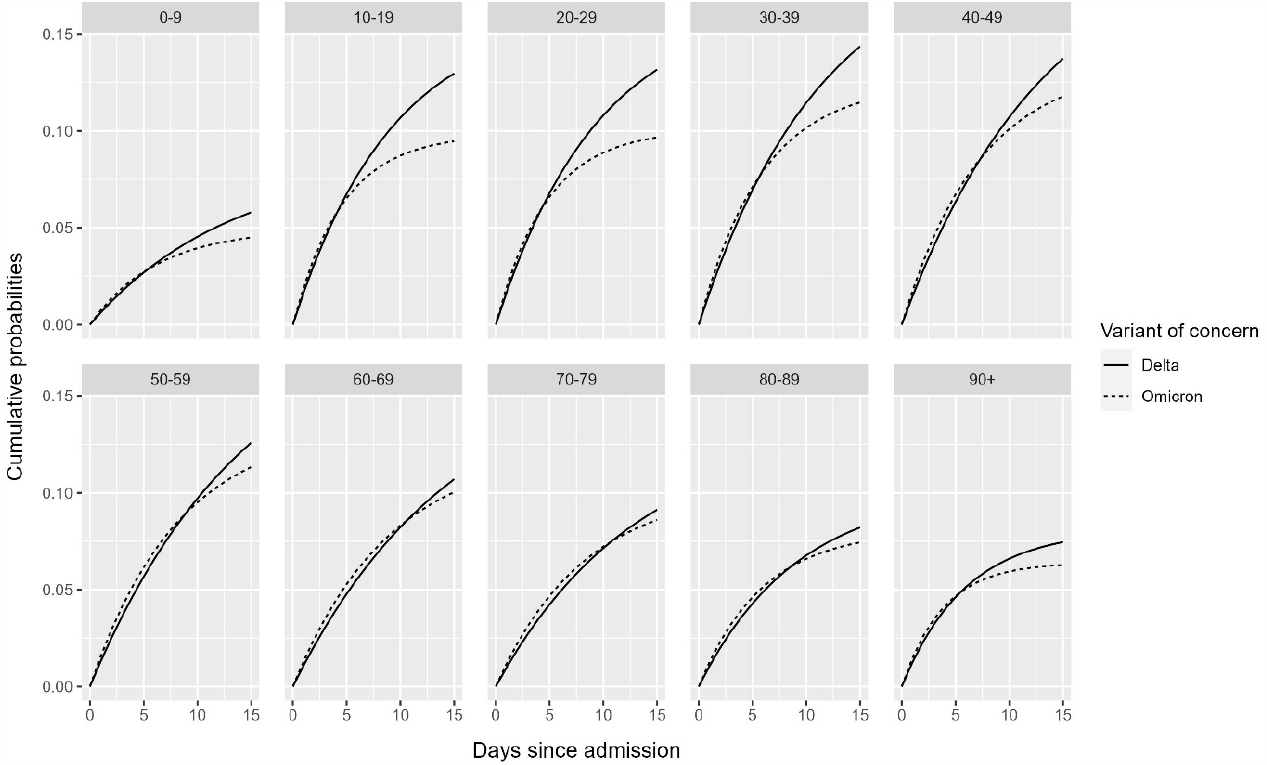
Estimated cumulative probabilities of transfer of COVID-19 patients in German intensive care units up to day 15 since admission, stratified by SARS-CoV-2 variant of concern infection and age group

## Results

### Transfer, discharge and death rates of COVID-19 patients in intensive care

During the selected calendar periods, a total of 6,046 transfers, 35,425 discharges and 12,114 deaths were observed. The multivariable Poisson regression analysis showed that Omicron infection was associated with higher transfer and discharge rates in COVID-19 ICU patients than Delta infection, with estimated adjusted IRR of 1.23 (95% CI, 1.16 – 1.30) and 2.27 (95% CI, 2.20 – 2.34), respectively (Table 1). With an estimated adjusted IRR of 0.98 (95% CI, 0.94 – 1.02), Delta and Omicron infection were both associated with nearly similar death rates.

Compared with the 60-69 age group, younger COVID-19 ICU patients were predominately associated with higher transfer rates, whereas older patients were predominantly associated with lower transfer rates (Table 1). For discharge, the estimated adjusted IRR peaked at 2.40 (95% CI, 2.15 – 2.67) in the 10-19 age group, then decreased steadily until the 60-69 age group and then increased again for older COVID-19 ICU patients. For death, younger age groups were associated with lower incidence rates compared with the 60-69 age group, whereas older COVID-19 ICU patients were associated with higher death rates, up to an estimated adjusted IRR of 4.09 (95% CI, 3.74 – 4.47) for the 90+ age group.

When comparing sex categories, male COVID-19 ICU patients were associated with lower transfer and discharge rates but with an estimated adjusted IRR of 1.04 (95% CI, 1.00 – 1.08) with a death rate nearly similar to that of female COVID-19 ICU patients (Table 1).

### Risk of transfer, discharge and death of COVID-19 patients in intensive care

The multistate analysis yielded a slightly higher estimated risk of transfer for COVID-19 ICU patients with Omicron infection at the beginning of observed ICU stay compared with those with Delta infection across all age groups (Fig. 2). However, the estimated risk of transfer associated with Omicron infection decreased with longer observed ICU stay and fell below the estimated risk of transfer associated with Delta infection, where estimated cumulative probabilities were still above 0.12 on day 15 after ICU admission at ages 10 to 59 years.

For discharge, estimated cumulative probabilities decreased across age groups (Fig. 3). For example, on day 10 after ICU admission, the estimated cumulative probability associated with Omicron infection was 0.77 for the 0-9 age group and 0.53 for 80-89 age group, respectively 0.48 (0-9 age group) and 0.29 (80-89 age group) with Delta infection. Thus, COVID-19 ICU patients with Omicron infection were consistently found to have a higher estimated risk of discharge compared with those with Delta infection across all age groups.

**Fig. 3.**
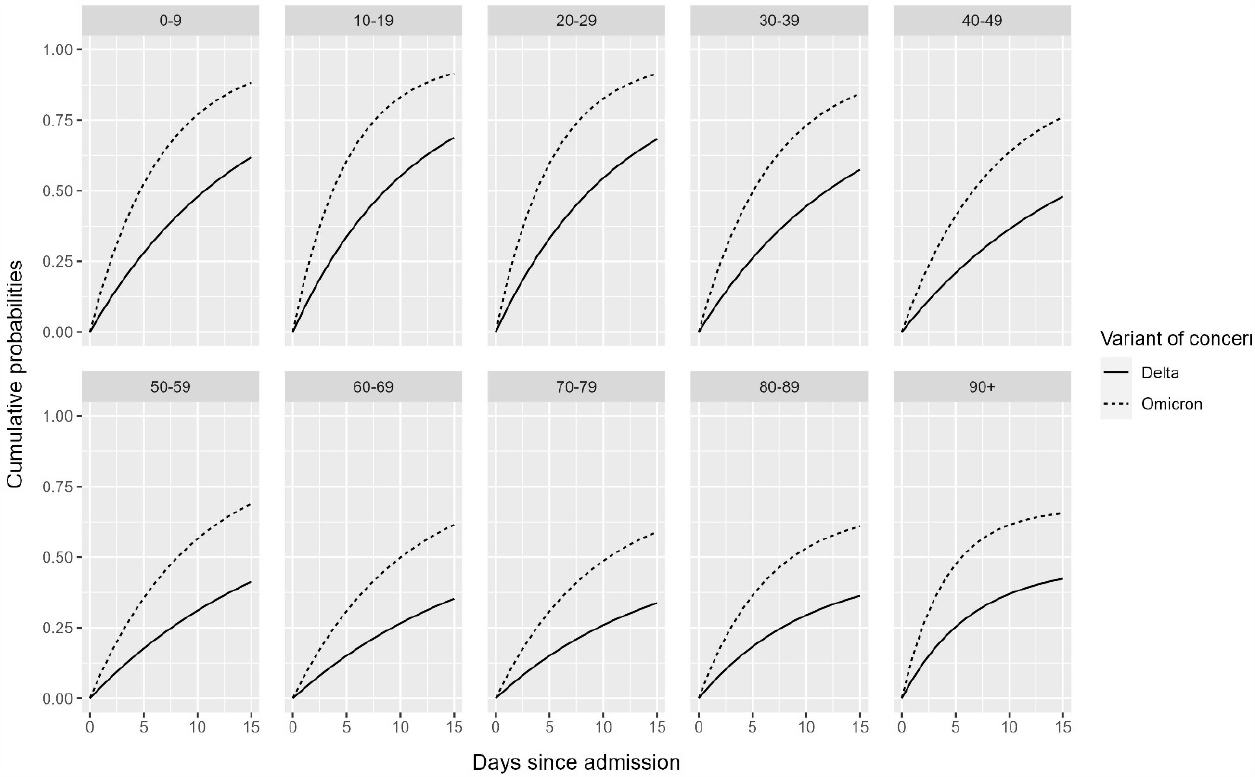
Estimated cumulative probabilities of discharge of COVID-19 patients in German intensive care units up to day 15 since admission, stratified by SARS-CoV-2 variant of concern infection and age group

For death, estimated cumulative probabilities increased sharply with age (Fig. 4). For example, the estimated cumulative probability on day 10 after ICU admission associated with Omicron infection was 0.01 for the 0-9 age group and 0.26 for the 80-89 age group, respectively 0.02 (0-9 age group) and 0.33 (80-89 age group) with Delta infection. Consequently, COVID-19 ICU patients with Omicron infection were consistently found to have a lower estimated risk of death compared with those with Delta infection across all age groups.

**Fig. 4.**
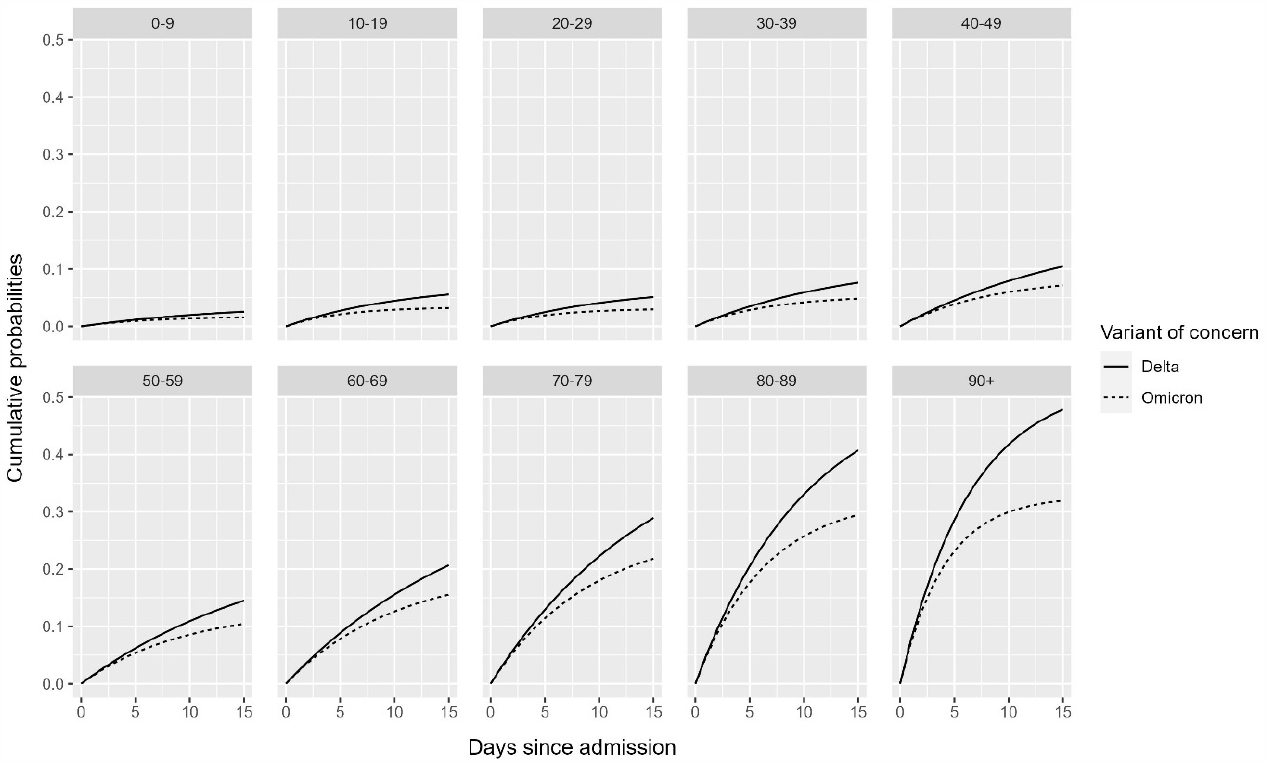
Estimated cumulative probabilities of death of COVID-19 patients in German intensive care units up to day 15 since admission, stratified by SARS-CoV-2 variant of concern infection and age group

## Discussion

In this retrospective cohort study, we aimed to determine the impact of Delta and Omicron VOC on critical care in Germany, particularly on inter-hospital transfers of COVID-19 ICU patients. Therefore, we performed a multivariable Poisson regression analysis to estimate the incidence rates and IRR of transfer, discharge, and death in COVID-19 ICU patients associated with Delta and Omicron infection. In addition, to estimate the associated risk of transfer, discharge and death, we applied a multistate model for the clinical trajectory of COVID-19 patients that accounts for these competing events during ICU stay. Our analysis was based on nationwide inpatient reimbursement data provided by the InEK and the calendar periods for Delta and Omicron VOC, which served as surrogate measures of infection impact, were selected using weekly sequence data on the number and proportion of VOC in Germany published by the RKI [13, 32].

### Transfer, discharge and death rates of COVID-19 patients in intensive care

The results of our multivariable Poisson regression analysis reveal the differential impact of the studied VOC on ICU stay of COVID-19 patients in Germany, for whom Omicron infection was associated with higher transfer and discharge rates compared with Delta infection. One plausible explanation might be comparatively more stable patients with a better prognosis, which made it possible to free up ICU capacities at all during periods of increased resource demand [39, 40]. Accordingly, the finding that ICU patients with Omicron infection were transferred and discharged at higher rates, thus more frequently, compared with those with Delta infection suggests that the severity of COVID-19 disease was also lower and confirms previous evidence [23, 24]. With respect to age, our findings show that older COVID-19 ICU patients were transferred and discharged less frequently, but died more frequently. This supports existing findings indicating the relevance of age, particularly in the context of COVID-19 related in-hospital mortality and the consequent need of close therapeutic care [36-38]. In addition, we found that male COVID-19 ICU patients were transferred and discharged less frequently compared to female COVID-19 ICU patients, suggesting worse clinical progressions [41, 42].

### Risk of transfer, discharge and death of COVID-19 patients in intensive care

The differential severity of COVID-19 disease associated with the studied VOC becomes even more apparent when considering the results of our multistate approach: ICU patients with Delta infection had a comparatively higher estimated risk of inter-hospital transfer than those with Omicron infection; across all age groups and the longer the observed ICU stay. In addition, the estimated risk of being discharged from ICU was found to be comparatively higher for COVID-19 patients with Omicron infection, whereas the estimated risk to die in ICU was higher for those with Delta infection. Hence, our findings support previous studies showing the reduced severity of COVID-19 infection with Omicron [24, 43-45]. Irrespective of predominant VOC, the influence of age became obvious and particularly so for the estimated risk of death in ICU, again indicating the close therapeutic demands in hospitalized elderly COVID-19 patients [36-38].

### Conversion of rates to risks in the presence of competing events

At first sight, one might suspect a contradiction when comparing our results on the rates and risks of the competing events of transfer, discharge and death. According to the multivariable Poisson regression analysis, Omicron infection was associated with comparatively higher transfer and discharge rates in COVID-19 ICU patients than Delta infection, whereas the associated death rates were almost the same for both VOC infections. However, converting rates to cumulative probabilities, i.e., risks, using the multistate model yielded comparatively lower estimated risk of transfer and death for COVID-19 ICU patients with Omicron infection across all age groups. This apparent contradiction is explained by the fact that rate and risk are different measures and, according to the multistate model, the estimated risk for each competing event *CIF*_01_(*t*), *CIF*_02_(*t*), *CIF*_03_ (*t*) depend on all corresponding hazard rates [46, 47]. For example, COVID-19 ICU patients with Omicron infection were transferred more frequently than those with Delta infection, with an estimated adjusted IRR above 1.0. However, when the competing event of discharge with an estimated adjusted IRR even further above 1.0 was also considered, this difference in transfer rates converted into a comparatively lower estimated risk of transfer for COVID-19 ICU patients with Omicron infection. Likewise, even though COVID-19 patients with Omicron infection died not less frequently in ICU than those with Delta infection, with an estimated adjusted IRR ∼1, their estimated risk of death was comparatively lower since they were discharged more frequently. Thus, the modelled clinical trajectory of COVID-19 ICU patients demonstrates the loss of the one-to-one correspondence between rate and risk, which is a distinctive feature of competing event scenarios [48].

### Strengths and Limitations

One of the strengths of the present study is the representativeness of the data source, which includes the remuneration data of all hospitals in Germany that are subject to the data transmission obligation to the InEK [32]. These data allowed us to gain further insight into critical care during the dynamic pandemic situation in Germany. In this context, our findings demonstrate differential severity of COVID-19 disease in ICU patients associated with Delta and Omicron infection and the related allocation of critically ill COVID-19 patients between hospitals. Moreover, the relevance of age for COVID-19 related in-hospital mortality was further substantiated [36-38]. From a methodological perspective, we have shown that the multistate approach is an appropriate means of modeling the state transitions of ICU patients in the presence of competing risks and using aggregate count data [30, 31, 33-35]. To our knowledge, there are no comparable approaches to address the rates and risks of inter-hospital transfers of COVID-19 ICU patients.

An important limitation with regard to the chosen methodology of our study are the underlying assumptions of constant hazard rates, independent discharge and death rates with respect to inter-hospital transfers, COVID-19 ICU patients were transferred only once during their inpatient treatment and discharges and deaths occurred exclusively in ICU. This might not reflect the actual course of clinical care. Second, we chose calendar periods as surrogate measures of the impact of Delta and Omicron infection, although other population-based measures, such as vaccination coverage or nonpharmacologic interventions, and patient characteristics may also have an influenced the situation in German ICU during these periods. However, these effects could not be separated by the data used. Adequate adjustment of the multistate model and more sophisticated analysis would therefore require information on patient-level.

## Conclusions

Comparison of the impact of Delta and Omicron VOC on critical care in Germany points toward a reduced severity of COVID-19 disease in ICU patients with Omicron infection, as they were transferred and discharged more frequently than those with Delta infection, who in turn had a substantially higher estimated risk of death. Age emerges as a relevant determinant for fatal clinical trajectories in COVID-19 ICU patients and imposes close therapeutic care.

## Data Availability

The data used in this study are available are available from the corresponding author on reasonable request but are subject to approval by the Federal Ministry of Health.

## Notes

### Competing Interest Statement

The authors have declared no competing interest.

### Funding Statement

All authors declare that no funds, grants, or other support were received for this study.

